# Analysis of exome-sequenced UK Biobank subjects implicates genes affecting risk of hyperlipidaemia

**DOI:** 10.1101/2020.07.09.20150334

**Authors:** David Curtis

## Abstract

Rare genetic variants in *LDLR, APOB* and *PCSK9* are known causes of familial hypercholesterolaemia and it is expected that rare variants in other genes will also have effects on hyperlipidaemia risk although such genes remain to be identified. The UK Biobank consists of a sample of 500,000 volunteers and exome sequence data is available for 50,000 of them. 11,490 of these were classified as hyperlipidaemia cases on the basis of having a relevant diagnosis recorded and/or taking lipid-lowering medication while the remaining 38,463 were treated as controls. Variants in each gene were assigned weights according to rarity and predicted impact and overall weighted burden scores were compared between cases and controls, including population principal components as covariates. One biologically plausible gene, *HUWE1*, produced statistically significant evidence for association after correction for testing 22,028 genes with a signed log10 p value (SLP) of −6.15, suggesting a protective effect of variants in this gene. Other genes with uncorrected p<0.001 are arguably also of interest, including *LDLR* (SLP=3.67), *RBP2* (SLP=3.14), *NPFFR1* (SLP=3.02) and *ACOT9* (SLP=-3.19). Gene set analysis indicated that rare variants in genes involved in metabolism and energy can influence hyperlipidaemia risk. Overall, the results provide some leads which might be followed up with functional studies and which could be tested in additional data sets as these become available. This research has been conducted using the UK Biobank Resource.

## Introduction

Hyperlipidaemia is an important risk factor for cardiovascular disease which is modified by genetic variation and some of the genes involved have been identified (Sharifi et al., 2017). Rare variants with a large dominant effect in *LDLR, APOB* and *PCSK9* cause 40% of cases of familial hypercholesterolaemia (FH), a severe form of hyperlipidaemia. A whole genome sequencing study of over 16,000 subjects failed to identify rare variants in any additional genes influencing lipid levels, though yielded a total of 26 loci containing common variants which were genome-wide significant, largely replicating findings from previous genome wide association studies (GWAS) (Natarajan et al., 2018; Willer et al., 2013). A GWAS of medication usage in 320,000 European UK Biobank participants identified 55 independent SNPs associated with taking statins, including 19 previously shown to be associated with low density lipoprotein cholesterol (LDLC), and showed that these were enriched in a number of gene sets involved with lipid metabolism (Wu et al., 2019). Another GWAS of European UK Biobank participants identified hundreds of genome-wide significant independent SNPs associated with lipid levels (Richardson et al., 2020).

Although common genetic variation makes a substantial overall contribution to the variance in lipid levels, selection pressures mean that common variants individually have small effects and it can be difficult to elucidate the biological mechanisms underlying their association. It is to be expected that rare variants will also have effects and it is possible that some of them might have large effect sizes. Such analyses may be better able to identify specific genes rather than genetic loci and may indicate a direction of effect. The UK Biobank sample (http://www.ukbiobank.ac.uk/about-biobank-uk/) contains information about medication usage and clinical diagnoses, both as reported by participants and as extracted from their health records. Exome sequence data is available for 50,000 subjects and a gene-wise weighted burden analysis of rare variants was carried out to attempt to identify genes associated with a hyperlipidaemia phenotype, defined as subjects with a diagnosis of hyperlipidaemia and/or taking cholesterol-lowering medication.

## Methods

The UK Biobank dataset was downloaded along with the variant call files for 49,953 subjects who had undergone exome-sequencing and genotyped using the GRCh38 assembly with coverage 20X at 94.6% of sites on average (Hout et al., 2019). UK Biobank had obtained ethics approval from the Research Ethics Committee (REC; approval number: 11/NW/0382) and informed consent from all participants. All variants were annotated using VEP, PolyPhen and SIFT (Adzhubei et al., 2013; Kumar et al., 2009; McLaren et al., 2016). To obtain population principal components reflecting ancestry, version 1.90beta of *plink* (https://www.cog-genomics.org/plink2) was run with the options *--maf 0*.*1 --pca header tabs --make-rel* (Chang et al., 2015; Purcell et al., 2007, 2009).

The hyperlipidaemia phenotype was determined from four sources in the dataset: self-reported high cholesterol; reporting taking cholesterol lowering medication; reporting taking a named statin; having an ICD10 diagnosis for hyperlipidaemia in hospital records or as a cause of death. Subjects in any of these categories were deemed to be cases with hyperlipidaemia while all other subjects were taken to be controls.

SCOREASSOC was used to carry out a weighted burden analysis to test whether, in each gene, sequence variants which were rarer and/or predicted to have more severe functional effects occurred more commonly in cases than controls. Attention was restricted to rare variants with minor allele frequency (MAF) <= 0.01. As previously described, variants were weighted by MAF so that variants with MAF=0.01 were given a weight of 1 while very rare variants with MAF close to zero were given a weight of 10 (Curtis, 2020). Variants were also weighted according to their functional annotation using the default weights provided with the GENEVARASSOC program, which was used to generate input files for weighted burden analysis by SCOREASSOC (Curtis, 2016, 2012). For example, a weight of 5 was assigned for a synonymous variant, 10 for a non-synonymous variant and 20 for a stop gained variant. Additionally, 10 was added to the weight if the PolyPhen annotation was possibly or probably damaging and also if the SIFT annotation was deleterious, meaning that a non-synonymous variant annotated as both damaging and deleterious would be assigned an overall weight of 30. The full set of weights is shown in Supplementary Table S1, copied from the previous reports which used this method (Curtis et al., 2019, 2018). The weight due to MAF and the weight due to functional annotation were then multiplied together to provide an overall weight for each variant. Variants were excluded if there were more than 10% of genotypes missing in the controls or if the heterozygote count was smaller than both homozygote counts in the controls. If a subject was not genotyped for a variant then they were assigned the subject-wise average score for that variant. For each subject a gene-wise weighted burden score was derived as the sum of the variant-wise weights, each multiplied by the number of alleles of the variant which the given subject possessed. For variants on the X chromosome, hemizygous males were treated as homozygotes.

For each gene, a ridge regression analysis was carried out with lamda=1 to test whether the gene-wise variant burden score was associated with the hyperlipidaemia phenotype. To do this, SCOREASSOC first calculates the likelihood for the phenotypes as predicted by the first 20 population principal components and then calculates the likelihood using a model which additionally incorporates the gene-wise burden scores. It then carries out a likelihood ratio test assuming that twice the natural log of the likelihood ratio follows a chi-squared distribution with one degree of freedom to produce a p value. The statistical significance is summarised as a signed log p value (SLP) which is the log base 10 of the p value given a positive sign if the score is higher in cases and negative if it is higher in controls. We have shown that incorporating population principal components in this way satisfactorily controls for test statistic inflation when applied to this heterogeneous dataset (Curtis, 2020).

Gene set analyses were carried out using the 1454 “all GO gene sets, gene symbols” pathways as listed in the file *c5*.*all*.*v5*.*0*.*symbols*.*gmt* downloaded from the Molecular Signatures Database at http://www.broadinstitute.org/gsea/msigdb/collections.jsp (Subramanian et al., 2005). For each set of genes, the natural logs of the gene-wise p values were summed according to Fisher’s method to produce a chi-squared statistic with degrees of freedom equal to twice the number of genes in the set. The p value associated with this chi-squared statistic was expressed as a minus log10 p (MLP) as a test of association of the set with the hyperlipidaemia phenotype.

## Results

There were 11,490 cases with a diagnosis of hyperlipidaemia and/or taking cholesterol-lowering medication and 38,463 controls. There were 22,028 genes for which there were qualifying variants and the QQ plot for the SLPs obtained for each gene is shown in Figure 1. This shows that the test is well-behaved and conforms well with the expected distribution.

**Figure 1.**
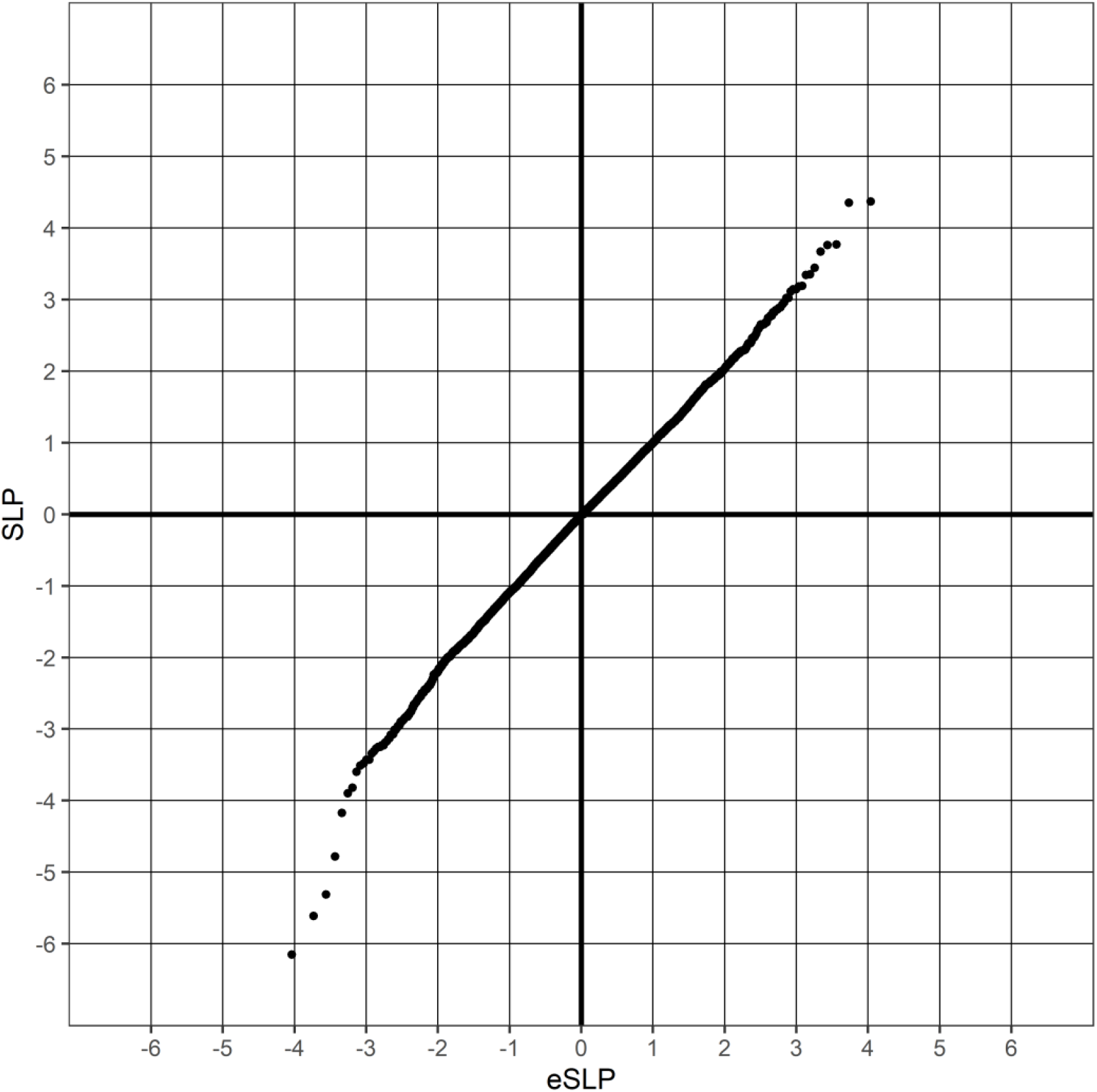
QQ plot of SLPs obtained for weighted burden analysis of 22,028 genes for association with hyperlipidaemia showing observed against expected SLP for each gene.

Table 1 shows the results for all genes with an absolute value of SLP exceeding 3 (equivalent to p<0.001). By chance, from 22,028 genes one would expect 11 to have SLP greater than 3 and 11 to have SLP less than −3, whereas the actual numbers are 15 and 28. Applying a Bonferroni correction to test for genome-wide statistical significance would yield a threshold of log10(22,028/0.05)=5.6 for the absolute value of the SLP and this is achieved by only two genes, *HUWE1* and *CXorf56*. For both these genes the SLP was negative, indicating that rare, functional variants were enriched in controls compared with cases and suggesting that impairment of these genes might be protective against hyperlipidaemia.

**Table 1.**
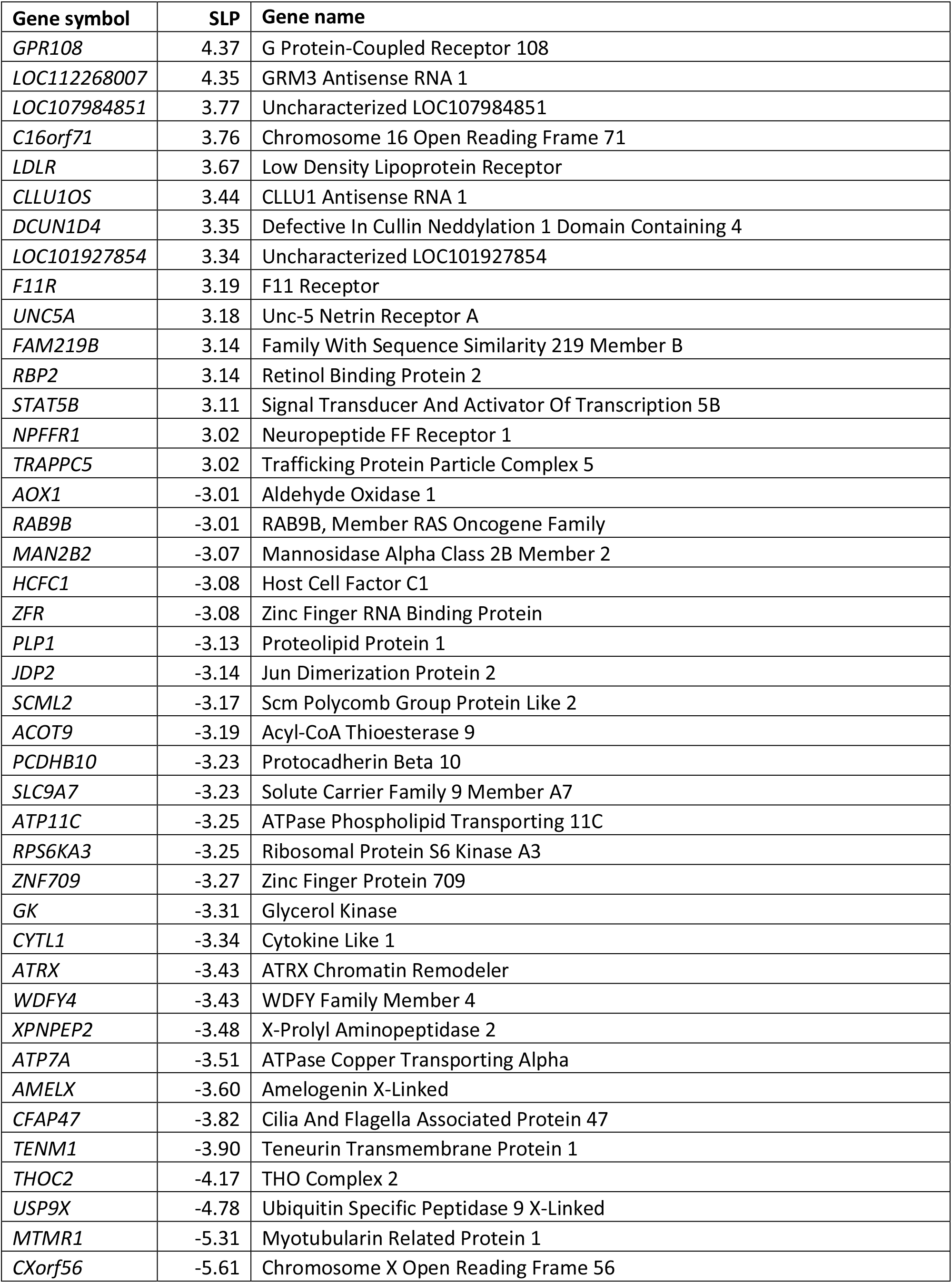
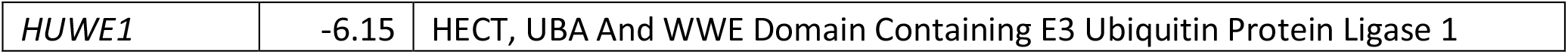
Genes with absolute value of SLP exceeding 3 or more (equivalent to p<0.001) for test of association of weighted burden score with hyperlipidaemia.

*HUWE1* (SLP=-6.15) codes for an ubiquitin protein ligase which has functions in development and tumorigenesis and which regulates the ABCG1 and ABCG4 lipid transporters such that over-expression of *HUWE1* reduces ABCG1 and ABCG4 protein levels and their cholesterol export activity while silencing the gene has the reverse effects (Aleidi et al., 2015). It is also mediates the ubiquitination of peroxisome proliferator-activated receptor α (PPARα), thereby reducing its function (Zhao et al., 2018). PPARα is a transcriptional factor which promotes hepatic lipid catabolism by stimulating fatty acid oxidation and ketogenesis in response to nutrient starvation.

*CXorf56* (SLP=-5.61) is a brain expressed gene which is not well characterised but has recently been implicated as a cause of intellectual disability (Rocha et al., 2020). Subjects with variants damaging this gene are reported to have intellectual disability and sometimes related features including epilepsy and alopecia but there are no reports of abnormalities of lipid metabolism.

No other gene was formally genome-wide significant and it is reasonable to assume that for most of those listed in the table the SLPs represent chance findings. However for a few it is possible that there is a real biological effect which is being picked up. The most obvious of these is *LDLR*, with SLP=3.67. It is well known that variants in this gene can cause familial hyperlipidaemia (Sharifi et al., 2017). It is worth noting however that the other two genes implicated in autosomal dominant familial hyperlipidaemia, *APOB* (SLP=0.11) and *PCSK9* (SLP=-0.66) did not produce any evidence of association. The same applies to *HMGCR* (SLP=-0.38), which codes for the target of statins.

Others of the genes with magnitude of SLP exceeding 3 (equivalent to p<0.001) which might be of interest are as follows.

*RBP2* (SLP=3.14) is expressed in the gut and its product is responsible for uptake of vitamin A but recent studies in mice have shown that it also influences body weight, the response to glucose challenge and hepatic triglyceride levels while *Rbp2* deficient mice have increased adiposity, with larger adipocytes, and decreased energy expenditure (Lee et al., 2020). The authors suggest a signalling role for RBP2 and if similar mechanisms were present in humans then it is plausible that variants disrupting *RBP2* could lead to increased risk of hyperlipidaemia.

*STAT5B* (SLP=3.11) codes for a transcription factor which is responsible for mediating the signal from various ligands, including growth hormone, and is also involved in adipogenesis (Goupille et al., 2016; Wakao et al., 2011). Two common variants in *STAT5B*, rs8082391 and rs8064638, have previously been reported to be associated with total cholesterol and low-density lipoprotein cholesterol while mice deficient in hepatic *Stat5a/b* had reduced serum cholesterol (Kornfeld et al., 2011). Additionally, a small candidate gene study claimed that SNPs in *STAT5B* were associated with lipid changes in response to growth hormone replacement therapy (Makimura et al., 2011).

However no SNPs close to *STAT5B* were genome-wide significantly associated with low density lipoprotein cholesterol or triglyceride levels in 180,000 UK Biobank subjects so the early SNP association results may have been false positives (Richardson et al., 2020). The mouse phenotype remains of interest, although it might suggest that variants in the gene should reduce, rather than increase, the risk of hyperlipidaemia

*NPFFR1* (SLP=3.02) codes for a receptor for a variety of neuropeptides including NPAF and NPFF and is expressed on adipocytes, where NPFF or NPAF treatment results in increased expression of adrenergic receptors and potentiation of the response to beta agonists to increase adenylyl cyclase activity (Lefrère et al., 2002). It is also expressed in the brain and NPFF has been shown to be anorexigenic in rats and chicks (Cline et al., 2007; Murase et al., 1996). Knockout of *Npffr1* in mice has differential effects according to sex including increased susceptibility to high fat diet with impaired glucose tolerance in males but increased weight and sensitivity to obesogenic insults in females (Leon et al., 2018). Overall it seems possible that impaired functioning of this receptor might lead to metabolic abnormalities which predispose to hyperlipidaemia.

*ACOT9* (SLP=-3.19) codes for a mitochondrial acyl-CoA thioesterase which has recently been proposed to have a key role in liver lipogenesis and risk of non-alcoholic fatty liver disease (NAFLD) (Steensels et al., 2020). It is a regulator of lipid accumulation and its expression is higher in NAFLD patients, while *Acot9* deficient mice are protected against weight gain, hepatic glucose production, steatosis and steatohepatitis in the setting of excess nutrition. These observations are consistent with the possibility that variants in *ACOT9* might tend to be protective against metabolic responses resulting in hyperlipidaemia.

*GK* (SLP=-3.31) codes for glycerol kinase and a number of variants in the gene have been reported to cause X-linked glycerol kinase deficiency syndromes which can very mild or which may involve symptoms including vomiting, hypoglycaemia, hyperketonaemia and intellectual disability (Sjarif et al., 1998). Its product has recently been shown to enhance hepatic liver metabolism and increased expression in mice leads to increased blood levels of cholesterol and triglycerides (Miao et al., 2020). Thus it is possible that variants mildly impairing *GK* but not sufficient to cause clinical glycerol kinase deficiency might be somewhat protective against hyperlipidaemia.

The gene sets with MLP>3 (equivalent to uncorrected p<0.001) are shown in Table 2. Given that 1454 sets were tested the critical value for the MLP to reach if a Bonferroni correction were applied would be log10(1454/0.05)=4.46, though this may be somewhat conservative given that some gene sets overlap with each other. This threshold is reached by the set HISTONE_MODIFICATION which contains 23 genes, one of which is *HUWE1* (SLP=-6.15). No other genes in the set appear likely to have a direct role in risk of hyperlipidaemia and it appears that the result for the set is essentially driven by this single gene. The other set to be formally statistically significant is GENERATION_OF_PRECURSOR_METABOLITES_AND_ENERGY with MLP=5.47, implying that variants disrupting the function of one or more genes in this set can impact hyperlipidaemia risk. The set contains 120 genes, none of which individually produced an SLP with magnitude greater than 3. However Table 3 shows the members of this set which have individual SLP>=1.3, equivalent to an uncorrected p value of 0.05, and a number of them appear to be of interest, as detailed below. These genes all have negative SLPs, suggesting that rare variants impacting their functioning might tend to be protective against hyperlipidaemia.

**Table 2.** Gene sets with value of MLP of 3 or more (equivalent to p<=0.01) for test of association of weighted burden scores with hyperlipidaemia.

| Gene set | MLP |
| --- | --- |
| GENERATION_OF_PRECURSOR_METABOLITES_AND_ENERGY | 5.47 |
| HISTONE_MODIFICATION | 4.46 |
| COVALENT_CHROMATIN_MODIFICATION | 4.24 |
| PROTEIN_SECRETION | 4.19 |
| HYDROGEN_ION_TRANSMEMBRANE_TRANSPORTER_ACTIVITY | 4.10 |
| MONOVALENT_INORGANIC_CATION_TRANSMEMBRANE_TRANSPORTER_ACTIVITY | 3.74 |
| ATPASE_ACTIVITY_COUPLED | 3.59 |
| SECRETION_BY_CELL | 3.48 |
| REGULATION_OF_PROTEIN_SECRETION | 3.34 |
| HYDROLASE_ACTIVITY_ACTING_ON_ACID_ANHYDRIDESCATALYZING_TRANSMEMBRANE_MOVEMENT_OF_SUBSTANCES | 3.24 |
| REGULATION_OF_CYTOKINE_SECRETION | 3.23 |
| SECRETION | 3.22 |
| ATPASE_ACTIVITY_COUPLED_TO_MOVEMENT_OF_SUBSTANCES | 3.22 |
| NERVOUS_SYSTEM_DEVELOPMENT | 3.17 |
| OXIDOREDUCTASE_ACTIVITY_ACTING_ON_THE_ALDEHYDE_OR_OXO_GROUP_OF_DONORS | 3.09 |
| LYASE_ACTIVITY | 3.08 |
| CYTOKINE_SECRETION | 3.05 |
| PURINE_RIBONUCLEOTIDE_BINDING | 3.05 |
| PRIMARY_ACTIVE_TRANSMEMBRANE_TRANSPORTER_ACTIVITY | 3.04 |
| MEMBRANE | 3.03 |

**Table 3.** Genes within GENERATION_OF_PRECURSOR_METABOLITES_AND_ENERGY with absolute value of SLP exceeding 1.3 (equivalent to p<0.05) for test of association of weighted burden score with hyperlipidaemia.

| Gene symbol | SLP | Gene name |
| --- | --- | --- |
| *PDHB* | 2.82 | Pyruvate Dehydrogenase E1 Subunit Beta |
| *ALDH5A1* | 2.38 | Aldehyde Dehydrogenase 5 Family Member A1 |
| *SLC25A3* | 1.62 | Solute Carrier Family 25 Member 3 |
| *ASIP* | 1.37 | Agouti Signaling Protein |
| *XYLB* | -1.31 | Xylulokinase |
| *ADIPOQ* | -1.37 | Adiponectin, C1Q And Collagen Domain Containing |
| *AVP* | -1.46 | Arginine Vasopressin |
| *SURF1* | -1.71 | SURF1 Cytochrome C Oxidase Assembly Factor |
| *ALDH9A1* | -1.72 | Aldehyde Dehydrogenase 9 Family Member A1 |
| *ADRB3* | -1.81 | Adrenoceptor Beta 3 |
| *ENOX2* | -1.83 | Ecto-NOX Disulfide-Thiol Exchanger 2 |
| *AKR1C3* | -1.84 | Aldo-Keto Reductase Family 1 Member C3 |
| *GLUD2* | -1.99 | Glutamate Dehydrogenase 2 |
| *GYG2* | -2.00 | Glycogenin 2 |
| *PHKA1* | -2.29 | Phosphorylase Kinase Regulatory Subunit Alpha 1 |
| *HCCS* | -2.56 | Holocytochrome C Synthase |
| *PHKA2* | -2.66 | Phosphorylase Kinase Regulatory Subunit Alpha 2 |

*ADIPOQ* (SLP=-1.37) codes for adiponectin, an adipokine which is expressed in adipocytes which influences fat metabolism, and common variants in the gene are associated with cardiovascular disease risk and lipid levels (Liu et al., 2018; Salazar-Tortosa et al., 2020; Wang et al., 2019; Zhang et al., 2019). Adiponectin has a hypoglycaemic effect and can reverse the insulin resistance associated with both lipoatrophy and obesity (Berg et al., 2001; Yamauchi et al., 2001). A rare intronic variant, rs74577862, has been reported to be associated with decreased levels of adiponectin and increased risk of atherosclerosis (Chen et al., 2017; UK10K Consortium et al., 2015).

*SURF1* (SLP=-1.71) codes for a protein involved in the assembly of cytochrome *c* oxidase complex and variants in it can cause Leigh syndrome, a fatal neurological condition (Lee et al., 2012). However mice completely lacking in *Surf1* have increased longevity, lower adiposity and enhanced fatty acid oxidation. These findings suggest that variants which moderately impact *SURF1* in humans might result in a more favourable lipid profile.

*ADRB3* (SLP=-1.81) codes for an adrenoceptor which promotes lipolysis and thermogenesis in response to sympathetic nerve stimulation and, as reviewed recently, the common variant Trp64Arg is associated with obesity and levels of serum lipids and adipokines (Luo et al., 2020).

*GYG2* (SLP=-2.00) has, like *SURF1*, been reported to be a cause of Leigh syndrome (Imagawa et al., 2014). Knockdown in adipocytes causes reduction in total lipid and number of lipid droplets per cell as well as increased lipolysis (Kerr et al., 2019).

*PHKA1* (SLP=-2.29) and *PHKA2* (SLP=-2.66) code for subunits of phosphorylase kinase and abnormalities in them are known to cause glycogen storage diseases with very variable phenotypes which can include hypoglycaemia, hypercholesterolaemia and hypertriglyceridaemia (Beauchamp et al., 2007; Preisler et al., 2012).

Inspection of the detailed results for all the genes mentioned revealed that the SLPs obtained tended to arise from the cumulative effects of many rare variants. For no gene was it possible to identify any individual variant which appeared to be a main driver for the overall differences in burden scores between cases and controls.

The SLPs for all genes and the MLPs for all gene sets are provided in supplementary tables S2 and S3.

## Discussion

This analysis identifies one gene, *HUWE1*, which reaches conventional standards for statistical significance after correction for multiple testing and which could plausibly have a role in influencing risk of hyperlipidaemia. An increased burden of rare functional variants is found in controls versus cases, implying that impaired functioning reduces hyperlipidaemia risk and suggesting that theoretically it might be considered a target for lipid-lowering pharmacotherapy, although one caveat is that it can act as either a tumour suppressor or as an oncogene (Crawford et al., 2020).

Other genes with uncorrected p values less than 0.001 are not formally statistically significant but might still be worth further investigation. The result for *LDLR* (SLP=3.67) demonstrates that even genes known to influence lipid levels may not achieve definitive results with a dataset of this size. Taking account of other information available, *RBP2* (SLP=3.14), *NPFFR1* (SLP=3.02) and *ACOT9* (SLP=-3.19) are especially noteworthy.

The significant results from the gene set analysis confirm that the variants tending to affect genes involved in metabolism and energy influence hyperlipidaemia risk but it is more difficult to say exactly which genes are involved. The results for *ADIPOQ* (SLP=-1.37) and *ADRB3* (SLP=-1.81) are in the opposite direction to what might be expected, since the literature would predict that disruption of these genes could promote hyperlipidaemia. On the other hand, it seems plausible that reduced function of *SURF1* (SLP=-1.71) or *GYG2* (SLP=-2.00) could be protective.

Overall, this analysis demonstrates that next generation sequence data does not provide a magic bullet for identifying rare variants influencing complex traits, even with large sample sizes. On the other hand, there are some potentially interesting results which are close to the threshold for formal significance such that it would be quite reasonable to expect that further definitive results could be obtained from a somewhat larger sample. At time of writing, exome sequencing of 200,000 UK Biobank samples has been completed and this data should be released soon and may well provide further insights.

## Data Availability

The raw data is available on application to UK Biobank. Detailed results with variant counts cannot be made available because they might be used for subject identification.

## Acknowledgments

This research has been conducted using the UK Biobank Resource. The author wishes to acknowledge the staff supporting the High Performance Computing Cluster, Computer Science Department, University College London. This work was carried out in part using resources provided by BBSRC equipment grant BB/R01356X/1.

---

**Supplementary Table 1.** The table shows the weight accorded to each type of variant as annotated by VEP (McLaren et al., 2016). 10 was added to this weight if the variant was annotated by Polyphen as possibly or probably damaging and 10 was added if SIFT annotated it as deleterious (Adzhubei et al., 2013; Kumar et al., 2009).

| VEP annotation | Weight |
| --- | --- |
| intergenic_variant | 1 |
| feature_truncation | 3 |
| regulatory_region_variant | 3 |
| feature_elongation | 3 |
| regulatory_region_amplification | 3 |
| regulatory_region_ablation | 3 |
| TF_binding_site_variant | 3 |
| TFBS_amplification | 3 |
| TFBS_ablation | 3 |
| downstream_gene_variant | 3 |
| upstream_gene_variant | 3 |
| non_coding_transcript_variant | 3 |
| NMD_transcript_variant | 3 |
| intron_variant | 3 |
| non_coding_transcript_exon_variant | 3 |
| 3_prime_UTR_variant | 5 |
| 5_prime_UTR_variant | 5 |
| mature_miRNA_variant | 5 |
| coding_sequence_variant | 5 |
| synonymous_variant | 5 |
| stop_retained_variant | 5 |
| incomplete_terminal_codon_variant | 5 |
| splice_region_variant | 5 |
| protein_altering_variant | 10 |
| missense_variant | 10 |
| inframe_deletion | 15 |
| inframe_insertion | 15 |
| transcript_amplification | 15 |
| start_lost | 15 |
| stop_lost | 15 |
| frameshift_variant | 20 |
| stop_gained | 20 |
| splice_donor_variant | 20 |
| splice_acceptor_variant | 20 |
| transcript_ablation | 20 |

## References

1. Adzhubei, I., Jordan, D.M., Sunyaev, S.R. (2013) Predicting functional effect of human missense mutations using PolyPhen-2. Curr. Protoc. Hum. Genet. 7 Unit7.20.

2. Aleidi, S.M., Howe, V., Sharpe, L.J., Yang, A., Rao, G., Brown, A.J., Gelissen, I.C. (2015) The E3 ubiquitin ligases, HUWE1 and NEDD4-1, are involved in the post-translational regulation of the ABCG1 and ABCG4 lipid transporters. J. Biol. Chem. 290, 24604–24613.

3. Beauchamp, N.J., Dalton, A., Ramaswami, U., Niinikoski, H., Mention, K., Kenny, P., Kolho, K.L., Raiman, J., Walter, J., Treacy, E., Tanner, S., Sharrard, M. (2007) Glycogen storage disease type IX: High variability in clinical phenotype. Mol. Genet. Metab. 92, 88–99.

4. Berg, A.H., Combs, T.P., Du, X., Brownlee, M., Scherer, P.E. (2001) The adipocyte-secreted protein Acrp30 enhances hepatic insulin action. Nat. Med. 7, 947–953.

5. Chang, C.C., Chow, C.C., Tellier, L.C., Vattikuti, S., Purcell, S.M., Lee, J.J. (2015) Second-generation PLINK: rising to the challenge of larger and richer datasets. Gigascience 4, 7.

6. Chen, X., Yuan, Y., Gao, Y., Wang, Q., Xie, F., Xia, D., Wei, Y., Xie, T. (2017) Association of variant in the ADIPOQ gene and functional study for its role in atherosclerosis. Oncotarget 8, 86527–86534.

7. Cline, M.A., Nandar, W., Rogers, J.O. (2007) Central neuropeptide FF reduces feed consumption and affects hypothalamic chemistry in chicks. Neuropeptides 41, 433–439.

8. Crawford, L.J., Campbell, D.C., Morgan, J.J., Lawson, M.A., Down, J.M., Chauhan, D., McAvera, R.M., Morris, T.C., Hamilton, C., Krishnan, A., Rajalingam, K., Chantry, A.D., Irvine, A.E. (2020) The E3 ligase HUWE1 inhibition as a therapeutic strategy to target MYC in multiple myeloma. Oncogene 39.

9. Curtis, D. (2012) A rapid method for combined analysis of common and rare variants at the level of a region, gene, or pathway. Adv Appl Bioinform Chem 5, 1–9.

10. Curtis, D. (2016) Pathway analysis of whole exome sequence data provides further support for the involvement of histone modification in the aetiology of schizophrenia. Psychiatr. Genet. 26, 223–7.

11. Curtis, D. (2020) Multiple linear regression allows weighted burden analysis of rare coding variants in an ethnically heterogeneous population. bioRxiv 2020.06.11.145938.

12. Curtis, D., Bakaya, K., Sharma, L., Bandyopadhay, S. (2019) Weighted burden analysis of exome-sequenced late onset Alzheimer’s cases and controls provides further evidence for involvement of PSEN1 and demonstrates protective role for variants in tyrosine phosphatase genes. Ann Hum Genet 84, 291–302.

13. Curtis, D., Coelewij, L., Liu, S.-H., Humphrey, J., Mott, R. (2018) Weighted Burden Analysis of Exome-Sequenced Case-Control Sample Implicates Synaptic Genes in Schizophrenia Aetiology. Behav. Genet. 43, 198–208.

14. Goupille, O., Penglong, T., Kadri, Z., Granger-Locatelli, M., Fucharoen, S., Maouche-Chrétien, L., Prost, S., Leboulch, P., Chrétien, S. (2016) Inhibition of the acetyl lysine-binding pocket of bromodomain and extraterminal domain proteins interferes with adipogenesis. Biochem. Biophys. Res. Commun. 472, 624–630.

15. Hout, C.V. Van, Tachmazidou, I., Backman, J.D., Hoffman, J.X., Ye, B., Pandey, A.K., Gonzaga-Jauregui, C., Khalid, S., Liu, D., Banerjee, N., Li, A.H., Colm, O., Marcketta, A., Staples, J., Schurmann, C., Hawes, A., Maxwell, E., Barnard, L., Lopez, A., Penn, J., Habegger, L., Blumenfeld, A.L., Yadav, A., Praveen, K., Jones, M., Salerno, W.J., Chung, W.K., Surakka, I., Willer, C.J., Hveem, K., Leader, J.B., Carey, D.J., Ledbetter, D.H., Collaboration, G.-R.D., Cardon, L., Yancopoulos, G.D., Economides, A., Coppola, G., Shuldiner, A.R., Balasubramanian, S., Cantor, M., Nelson, M.R., Whittaker, J., Reid, J.G., Marchini, J., Overton, J.D., Scott, R.A., Abecasis, G., Yerges-Armstrong, L., Baras, A., Center, on behalf of the R.G. (2019) Whole exome sequencing and characterization of coding variation in 49,960 individuals in the UK Biobank. bioRxiv 572347.

16. Imagawa, E., Osaka, H., Yamashita, A., Shiina, M., Takahashi, E., Sugie, H., Nakashima, M., Tsurusaki, Y., Saitsu, H., Ogata, K., Matsumoto, N., Miyake, N. (2014) A hemizygous GYG2 mutation and Leigh syndrome: A possible link? Hum. Genet. 133, 225–234.

17. Kerr, A.G., Sinha, I., Dadvar, S., Arner, P., Dahlman, I. (2019) Epigenetic regulation of diabetogenic adipose morphology. Mol. Metab. 25, 159–167.

18. Kornfeld, J.-W., Isaacs, A., Vitart, V., Pospisilik, J.A., Meitinger, T., Gyllensten, U., Wilson, J.F., Rudan, I., Campbell, H., Penninger, J.M., Sexl, V., Moriggl, R., van Duijn, C., Pramstaller, P.P., Hicks, A.A. (2011) Variants in STAT5B Associate with Serum TC and LDL-C Levels. J. Clin. Endocrinol. Metab. 96, E1496–E1501.

19. Kumar, P., Henikoff, S., Ng, P.C. (2009) Predicting the effects of coding non-synonymous variants on protein function using the SIFT algorithm. Nat. Protoc. 4, 1073–1081.

20. Lee, I.C., El-Hattab, A.W., Wang, J., Li, F.Y., Weng, S.W., Craigen, W.J., Wong, L.J.C. (2012) SURF1-associated leigh syndrome: A case series and novel mutations. Hum. Mutat. 33, 1192–1200.

21. Lee, S.A., Zhang Yang, K.J., Brun, P.J., Silvaroli, J.A., Yuen, J.J., Shmarakov, I., Jiang, H., Feranil, J.B., Li, X., Lackey, A.I., Krężel, W., Leibel, R.L., Libien, J., Storch, J., Golczak, M., Blaner, W.S. (2020) Retinol-binding protein 2 (RBP2) binds monoacylglycerols and modulates gut endocrine signaling and body weight. Sci. Adv. 6.

22. Lefrère, I., De Coppet, P., Camelin, J.C., Lay, S. Le, Mercier, N., Elshourbagy, N., Bril, A., Berrebi-Bertrand, I., Fève, B., Krief, S. (2002) Neuropeptide AF and FF modulation of adipocyte metabolism. Primary insights from functional genomics and effects on β-adrenergic responsiveness. J. Biol. Chem. 277, 39169–39178.

23. Leon, S., Velasco, I., Vázquez, M.J., Barroso, A., Beiroa, D., Heras, V., Ruiz-Pino, F., Manfredi-Lozano, M., Romero-Ruiz, A., Sanchez-Garrido, M.A., Dieguez, C., Pinilla, L., Roa, J., Nogueiras, R., Tena-Sempere, M. (2018) Sex-Biased Physiological Roles of NPFF1R, the Canonical Receptor of RFRP-3, in Food Intake and Metabolic Homeostasis Revealed by its Congenital Ablation in mice. Metabolism. 87, 87–97.

24. Liu, Yawen, Kanu, J.S., Qiu, S., Cheng, Y., Li, R., Kou, C., Gu, Y., Bai, Y., Shi, J., Li, Y., Liu, Yunkai, Yu, Y. (2018) Associations between three common single nucleotide polymorphisms (rs266729, rs2241766, and rs1501299) of ADIPOQ and cardiovascular disease: A meta-analysis. Lipids Health Dis. 17.

25. Luo, Z., Zhang, T., Wang, S., He, Y., Ye, Q., Cao, W. (2020) The Trp64Arg polymorphism in β3 adrenergic receptor (ADRB3) gene is associated with adipokines and plasma lipids: A systematic review, meta-analysis, and meta-regression. Lipids Health Dis. 19.

26. Makimura, M., Ihara, K., Kojima-Ishii, K., Nozaki, T., Ohkubo, K., Kohno, H., Kishimoto, J., Hara, T. (2011) The signal transducer and activator of transcription 5B gene polymorphism contributes to the cholesterol metabolism in Japanese children with growth hormone deficiency. Clin. Endocrinol. (Oxf). 74, 611–617.

27. McLaren, W., Gil, L., Hunt, S.E., Riat, H.S., Ritchie, G.R.S., Thormann, A., Flicek, P., Cunningham, F. (2016) The Ensembl Variant Effect Predictor. Genome Biol. 17, 122.

28. Miao, L., Su, F., Yang, Y., Liu, Y., Wang, L., Zhan, Y., Yin, R., Yu, M., Li, C., Yang, X., Ge, C. (2020) Glycerol kinase enhances hepatic lipid metabolism by repressing nuclear receptor subfamily 4 group a1 in the nucleus. Biochem. Cell Biol. 98, 370–377.

29. Murase, T., Arima, H., Kondo, K., Oiso, Y. (1996) Neuropeptide FF reduces food intake in rats. Peptides 17, 353–354.

30. Natarajan, P., Peloso, G.M., Zekavat, S.M., Montasser, M., Ganna, A., Chaffin, M., Khera, A. V., Zhou, W., Bloom, J.M., Engreitz, J.M., Ernst, J., O’Connell, J.R., Ruotsalainen, S.E., Alver, M., Manichaikul, A., Johnson, W.C., Perry, J.A., Poterba, T., Seed, C., Surakka, I.L., Esko, T., Ripatti, S., Salomaa, V., Correa, A., Vasan, R.S., Kellis, M., Neale, B.M., Lander, E.S., Abecasis, G., Mitchell, B., Rich, S.S., Wilson, J.G., Cupples, L.A., Rotter, J.I., Willer, C.J., Kathiresan, S., Abe, N., Albert, C., Allred, N. (Nichole) P., Almasy, L., Alonso, A., Ament, S., Anderson, P., Anugu, P., Applebaum-Bowden, D., Arking, D., Arnett, D.K., Ashley-Koch, A., Aslibekyan, S., Assimes, T., Auer, P., Avramopoulos, D., Barnard, J., Barnes, K., Barr, R.G., Barron-Casella, E., Beaty, T., Becker, D., Becker, L., Beer, R., Begum, F., Beitelshees, A., Benjamin, E., Bezerra, M., Bielak, L., Bis, J., Blackwell, T., Blangero, J., Boerwinkle, E., Borecki, I., Bowler, R., Brody, J., Broeckel, U., Broome, J., Bunting, K., Burchard, E., Cardwell, J., Carty, C., Casaburi, R., Casella, J., Chang, C., Chasman, D., Chavan, S., Chen, B.J., Chen, W.M., Chen, Y.D.I., Cho, M., Choi, S.H., Chuang, L.M., Chung, M., Cornell, E., Crandall, C., Crapo, J., Curran, J., Curtis, J., Custer, B., Damcott, C., Darbar, D., Das, S., David, S., Davis, C., Daya, M., de Andrade, M., DeBaun, M., Deka, R., DeMeo, D., Devine, S., Do, R., Duan, Q., Duggirala, R., Durda, P., Dutcher, S., Eaton, C., Ekunwe, L., Ellinor, P., Emery, L., Farber, C., Farnam, L., Fingerlin, T., Flickinger, M., Fornage, M., Franceschini, N., Fu, M., Fullerton, M., Fulton, L., Gabriel, S., Gan, W., Gao, Y., Gass, M., Gelb, B., Geng, X. (Priscilla), Germer, S., Gignoux, C., Gladwin, M., Glahn, D., Gogarten, S., Gong, D.W., Goring, H., Gu, C.C., Guan, Y., Guo, X., Haessler, J., Hall, M., Harris, D., Hawley, N., He, J., Heavner, B., Heckbert, S., Hernandez, R., Herrington, D., Hersh, C., Hidalgo, B., Hixson, J., Hokanson, J., Hong, E., Hoth, K., Hsiung, C. (Agnes), Huston, H., Hwu, C.M., Irvin, M.R., Jackson, R., Jain, D., Jaquish, C., Jhun, M.A., Johnsen, J., Johnson, A., Johnston, R., Jones, K., Kang, H.M., Kaplan, R., Kardia, S., Kaufman, L., Kelly, S., Kenny, E., Kessler, M., Khan, A., Kinney, G., Konkle, B., Kooperberg, C., Kramer, H., Krauter, S., Lange, C., Lange, E., Lange, L., Laurie, Cathy, Laurie, Cecelia, LeBoff, M., Lee, S.S., Lee, W.J., LeFaive, J., Levine, D., Levy, D., Lewis, J., Li, Y., Lin, H., Lin, K.H., Liu, S., Liu, Y., Loos, R., Lubitz, S., Lunetta, K., Luo, J., Mahaney, M., Make, B., Manson, J.A., Margolin, L., Martin, L., Mathai, S., Mathias, R., McArdle, P., McDonald, M.L., McFarland, S., McGarvey, S., Mei, H., Meyers, D.A., Mikulla, J., Min, N., Minear, M., Minster, R.L., Musani, S., Mwasongwe, S., Mychaleckyj, J.C., Nadkarni, G., Naik, R., Nekhai, S., Nickerson, D., North, K., O’Connor, T., Ochs-Balcom, H., Pankow, J., Papanicolaou, G., Parker, M., Parsa, A., Penchev, S., Peralta, J.M., Perez, M., Peters, U., Peyser, P., Phillips, L., Phillips, S., Pollin, T., Post, W., Becker, J.P., Boorgula, M.P., Preuss, M., Prokopenko, D., Psaty, B., Qasba, P., Qiao, D., Qin, Z., Rafaels, N., Raffield, L., Rao, D.C., Rasmussen-Torvik, L., Ratan, A., Redline, S., Reed, R., Regan, E., Reiner, A., Rice, K., Roden, D., Roselli, C., Ruczinski, I., Russell, P., Ruuska, S., Ryan, K., Sakornsakolpat, P., Salimi, S., Salzberg, S., Sandow, K., Sankaran, V., Schmidt, E., Schwander, K., Schwartz, D., Sciurba, F., Seidman, C., Sheehan, V., Shetty, Amol, Shetty, Aniket, Sheu, W.H.H., Shoemaker, M.B., Silver, B., Silverman, E., Smith, Jennifer, Smith, Josh, Smith, N., Smith, T., Smoller, S., Snively, B., Sofer, T., Sotoodehnia, N., Stilp, A., Streeten, E., Sung, Y.J., Sylvia, J., Szpiro, A., Sztalryd, C., Taliun, D., Tang, H., Taub, M., Taylor, K., Taylor, S., Telen, M., Thornton, T.A., Tinker, L., Tirschwell, D., Tiwari, H., Tracy, R., Tsai, M., Vaidya, D., VandeHaar, P., Vrieze, S., Walker, T., Wallace, R., Walts, A., Wan, E., Wang, F.F., Watson, K., Weeks, D.E., Weir, B., Weiss, S., Weng, L.C., Willer, C., Williams, K., Williams, L.K., Wilson, C., Wong, Q., Xu, H., Yanek, L., Yang, I., Yang, R., Zaghloul, N., Zhang, Y., Zhao, S.X., Zheng, X., Zhi, D., Zhou, X., Zody, M., Zoellner, S. (2018) Deep-coverage whole genome sequences and blood lipids among 16,324 individuals. Nat. Commun. 9, 1–12.

31. Preisler, N., Ørngreen, M.C., Echaniz-Laguna, A., Laforet, P., Lonsdorfer-Wolf, E., Doutreleau, S., Geny, B., Akman, H.O., DiMauro, S., Vissing, J. (2012) Muscle phosphorylase kinase deficiency: A neutral metabolic variant or a disease? Neurology 78, 265–268.

32. Purcell, S., Neale, B., Todd-Brown, K., Thomas, L., Ferreira, M.A.R., Bender, D., Maller, J., Sklar, P., de Bakker, P.I.W., Daly, M.J., Sham, P.C. (2007) PLINK: a tool set for whole-genome association and population-based linkage analyses. Am. J. Hum. Genet. 81, 559–75.

33. Purcell, S.M., Wray, N.R., Stone, J.L., Visscher, P.M., O’Donovan, M.C., Sullivan, P.F., Sklar, P., Purcell Leader, S.M., Ruderfer, D.M., McQuillin, A., Morris, D.W., O’Dushlaine, C.T., Corvin, A., Holmans, P. a, Macgregor, S., Gurling, H., Blackwood, D.H.R., Craddock, N.J., Gill, M., Hultman, C.M., Kirov, G.K., Lichtenstein, P., Muir, W.J., Owen, M.J., Pato, C.N., Scolnick, E.M., St Clair, D., Sklar Leader, P., Williams, N.M., Georgieva, L., Nikolov, I., Norton, N., Williams, H., Toncheva, D., Milanova, V., Thelander, E.F., Sullivan, P.F., Kenny, E., Quinn, E.M., Choudhury, K., Datta, S., Pimm, J., Thirumalai, S., Puri, V., Krasucki, R., Lawrence, J., Quested, D., Bass, N., Crombie, C., Fraser, G., Leh Kuan, S., Walker, N., McGhee, K. a, Pickard, B., Malloy, P., Maclean, A.W., Van Beck, M., Pato, M.T., Medeiros, H., Middleton, F., Carvalho, C., Morley, C., Fanous, A., Conti, D., Knowles, J. a, Paz Ferreira, C., Macedo, A., Helena Azevedo, M., Kirby, A.N., Ferreira, M. a R., Daly, M.J., Chambert, K., Kuruvilla, F., Gabriel, S.B., Ardlie, K., Moran, J.L. (2009) Common polygenic variation contributes to risk of schizophrenia and bipolar disorder. Nature 10, 8192–8192.

34. Richardson, T.G., Sanderson, E., Palmerid, T.M., Korpelaid, M.A., Ference, B.A., Smith, G.D., Holmes, M. V. (2020) Evaluating the relationship between circulating lipoprotein lipids and apolipoproteins with risk of coronary heart disease: A multivariable Mendelian randomisation analysis. PLoS Med. 17, e1003062.

35. Rocha, M.E., Silveira, T.R.D., Sasaki, E., Sás, D.M., Lourenço, C.M., Kandaswamy, K.K., Beetz, C., Rolfs, A., Bauer, P., Reardon, W., Bertoli-Avella, A.M. (2020) Novel clinical and genetic insight into CXorf56-associated intellectual disability. Eur. J. Hum. Genet. 28, 367–372.

36. Salazar-Tortosa, D.F., Pascual-Gamarra, J.M., Labayen, I., Rupérez, A.I., Censi, L., Béghin, L., Michels, N., González-Gross, M., Manios, Y., Lambrinou, C.-P., Moreno, L.A., Meirhaeghe, A., Castillo, M.J., Ruiz, J.R. (2020) Single nucleotide polymorphisms of ADIPOQ gene associated with cardiovascular disease risk factors in European adolescents. J. Hypertens.

37. Sharifi, M., Futema, M., Nair, D., Humphries, S.E. (2017) Genetic Architecture of Familial Hypercholesterolaemia. Curr. Cardiol. Rep.

38. Sjarif, D.R., Sinke, R.J., Duran, M., Beemer, F.A., Kleijer, W.J., Van Ploos Amstel, J.K., Poll-The, B.T. (1998) Clinical heterogeneity and novel mutations in the glycerol kinase gene in three families with isolated glycerol kinase deficiency. J. Med. Genet. 35, 650–656.

39. Steensels, S., Qiao, J., Zhang, Y., Maner-Smith, K.M., Kika, N., Holman, C.D., Corey, K.E., Bracken, W.C., Ortlund, E.A., Ersoy, B.A. (2020) Acot9 traffics mitochondrial short-chain fatty acids towards de novo lipogenesis and glucose production in the liver. Hepatology.

40. Subramanian, A., Tamayo, P., Mootha, V.K., Mukherjee, S., Ebert, B.L., Gillette, M.A., Paulovich, A., Pomeroy, S.L., Golub, T.R., Lander, E.S., Mesirov, J.P. (2005) Gene set enrichment analysis: a knowledge-based approach for interpreting genome-wide expression profiles. Proc Natl Acad Sci U S A 102, 15545–15550.

41. UK10K Consortium, K., Walter, K., Min, J.L., Huang, J., Crooks, L., Memari, Y., McCarthy, S., Perry, J.R.B., Xu, C., Futema, M., Lawson, D., Iotchkova, V., Schiffels, S., Hendricks, A.E., Danecek, P., Li, R., Floyd, J., Wain, L. V, Barroso, Inês, Humphries, S.E., Hurles, M.E., Zeggini, E., Barrett, J.C., Plagnol, V., Richards, J.B., Greenwood, C.M.T., Timpson, N.J., Durbin, R., Soranzo, N., Clapham, P., Coates, G., Cox, T., Daly, A., Danecek, P., Du, Y., Durbin, R., Edkins, S., Ellis, P., Flicek, P., Guo, Xiaosen, Guo, Xueqin, Huang, L., Jackson, D.K., Joyce, C., Keane, T., Kolb-Kokocinski, A., Langford, C., Li, Y., Liang, J., Lin, H., Liu, R., Maslen, J., McCarthy, S., Muddyman, D., Quail, M.A., Stalker, J., Sun, J., Tian, J., Wang, G., Wang, J., Wang, Y., Wong, K., Zhang, P., Barroso In?s, Birney E., Boustred, C., Chen, L., Clement, G., Cocca, M., Danecek, P., Davey Smith, G., Day, I.N.M., Day-Williams, A., Down, T., Dunham, I., Durbin, R., Evans, D.M., Gaunt, T.R., Geihs, M., Greenwood, C.M.T., Hart, D., Hendricks, A.E., Howie, B., Huang, J., Hubbard, T., Hysi, P., Iotchkova, V., Jamshidi, Y., Karczewski, K.J., Kemp, J.P., Lachance, G., Lawson, D., Lek, M., Lopes, M., MacArthur, D.G., Marchini, J., Mangino, M., Mathieson, I., McCarthy, S., Memari, Y., Metrustry, S., Min, J.L., Moayyeri, A., Muddyman, D., Northstone, K., Panoutsopoulou, K., Paternoster, L., Perry, J.R.B., Quaye, L., Brent Richards, J., Ring, S., Ritchie, G.R.S., Schiffels, S., Shihab, H.A., Shin, S.-Y., Small, K.S., Soler Artigas, M., Soranzo, N., Southam, L., Spector, T.D., St Pourcain, B., Surdulescu, G., Tachmazidou, I., Timpson, N.J., Tobin, M.D., Valdes, A.M., Visscher, P.M., Wain, L. V., Walter, K., Ward, K., Wilson, S.G., Wong, K., Yang, J., Zeggini, E., Zhang, F., Zheng, H.-F., Anney, R., Ayub, M., Barrett, J.C., Blackwood, D., Bolton, P.F., Breen, G., Collier, D.A., Craddock, N., Crooks, L., Curran, S., Curtis, D., Durbin, R., Gallagher, L., Geschwind, D., Gurling, H., Holmans, P., Lee, I., L?nnqvist, J., McCarthy, S., McGuffin, P., McIntosh, A.M., McKechanie, A.G., McQuillin, A., Morris, J., Muddyman, D., O’Donovan, M.C., Owen, M.J., Palotie, A., Parr, J.R., Paunio, T., Pietilainen, O., Rehnstr?m, K., Sharp, S.I., Skuse, D., St Clair, D., Suvisaari, J., Walters, J.T.R., Williams, H.J., Barroso, In?s, Bochukova, E., Bounds, R., Dominiczak, A., Durbin, R., Farooqi, I.S., Hendricks, A.E., Keogh, J., Marenne, G., McCarthy, S., Morris, A., Muddyman, D., O’Rahilly, S., Porteous, D.J., Smith, B.H., Tachmazidou, I., Wheeler, E., Zeggini, E., Al Turki, S., Anderson, C.A., Antony, D., Barroso, In?s Beales P., Bentham, J., Bhattacharya, S., Calissano, M., Carss, K., Chatterjee, K., Cirak, S., Cosgrove, C., Durbin, R., Fitzpatrick, D.R., Floyd, J., Reghan Foley, A., Franklin, C.S., Futema, M., Grozeva, D., Humphries, S.E., Hurles, M.E., McCarthy, S., Mitchison, H.M., Muddyman, D., Muntoni, F., O’Rahilly, S., Onoufriadis, A., Parker, V., Payne, F., Plagnol, V., Lucy Raymond, F., Roberts, N., Savage, D.B., Scambler, P., Schmidts, M., Schoenmakers, N., Semple, R.K., Serra, E., Spasic-Boskovic, O., Stevens, E., van Kogelenberg, M., Vijayarangakannan, P., Walter, K., Williamson, K.A., Wilson, C., Whyte, T., Ciampi, A., Greenwood, C.M.T., Hendricks, A.E., Li, R., Metrustry, S., Oualkacha, K., Tachmazidou, I., Xu, C., Zeggini, E., Bobrow, M., Bolton, P.F., Durbin, R., Fitzpatrick, D.R., Griffin, H., Hurles, M.E., Kaye, J., Kennedy, K., Kent, A., Muddyman, D., Muntoni, F., Lucy Raymond, F., Semple, R.K., Smee, C., Spector, T.D., Timpson, N.J., Charlton, R., Ekong, R., Futema, M., Humphries, S.E., Khawaja, F., Lopes, L.R., Migone, N., Payne, S.J., Plagnol, V., Pollitt, R.C., Povey, S., Ridout, C.K., Robinson, R.L., Scott, R.H., Shaw, A., Syrris, P., Taylor, R., Vandersteen, A.M., Barrett, J.C., Barroso, In?s, Davey Smith, G., Durbin, R., Farooqi, I.S., Fitzpatrick, D.R., Hurles, M.E., Kaye, J., Kennedy, K., Langford, C., McCarthy, S., Muddyman, D., Owen, M.J., Palotie, A., Brent Richards, J., Soranzo, N., Spector, T.D., Stalker, J., Timpson, N.J., Zeggini, E., Amuzu, A., Pablo Casas, J., Chambers, J.C., Cocca, M., Dedoussis, G., Gambaro, G., Gasparini, P., Gaunt, T.R., Huang, J., Iotchkova, V., Isaacs, A., Johnson, J., Kleber, M.E., Kooner, J.S., Langenberg, C., Luan, J., Malerba, G., M?rz, W., Matchan, A., Min, J.L., Morris, R., Nordestgaard, B.G., Benn, M., Ring, S., Scott, R.A., Soranzo, N., Southam, L., Timpson, N.J., Toniolo, D., Traglia, M., Tybjaerg-Hansen, A., van Duijn, C.M., van Leeuwen, E.M., Varbo, A., Whincup, P., Zaza, G., Zeggini, E., Zhang, W. (2015) The UK10K project identifies rare variants in health and disease. Nature 526, 82–90.

42. Wakao, H., Wakao, R., Oda, A., Fujita, H. (2011) Constitutively active Stat5A and Stat5B promote adipogenesis. Environ. Health Prev. Med. 16, 247–252.

43. Wang, Z., Diao, J., Yue, X., Zhong, J. (2019) Effects of ADIPOQ polymorphisms on individual susceptibility to coronary artery disease: a meta-analysis. Adipocyte 8, 137–143.

44. Willer, C.J., Schmidt, E.M., Sengupta, S., Peloso, G.M., Gustafsson, S., Kanoni, S., Ganna, A., Chen, J., Buchkovich, M.L., Mora, S., Beckmann, J.S., Bragg-Gresham, J.L., Chang, H.Y., Demirkan, A., Den Hertog, H.M., Do, R., Donnelly, L.A., Ehret, G.B., Esko, T., Feitosa, M.F., Ferreira, T., Fischer, K., Fontanillas, P., Fraser, R.M., Freitag, D.F., Gurdasani, D., Heikkilä, K., Hyppönen, E., Isaacs, A., Jackson, A.U., Johansson, Å., Johnson, T., Kaakinen, M., Kettunen, J., Kleber, M.E., Li, X., Luan, J., Lyytikäinen, L.P., Magnusson, P.K.E., Mangino, M., Mihailov, E., Montasser, M.E., Müller-Nurasyid, M., Nolte, I.M., O’Connell, J.R., Palmer, C.D., Perola, M., Petersen, A.K., Sanna, S., Saxena, R., Service, S.K., Shah, S., Shungin, D., Sidore, C., Song, C., Strawbridge, R.J., Surakka, I., Tanaka, T., Teslovich, T.M., Thorleifsson, G., Van Den Herik, E.G., Voight, B.F., Volcik, K.A., Waite, L.L., Wong, A., Wu, Y., Zhang, W., Absher, D., Asiki, G., Barroso, I., Been, L.F., Bolton, J.L., Bonnycastle, L.L., Brambilla, P., Burnett, M.S., Cesana, G., Dimitriou, M., Doney, A.S.F., Döring, A., Elliott, P., Epstein, S.E., Eyjolfsson, G.I., Gigante, B., Goodarzi, M.O., Grallert, H., Gravito, M.L., Groves, C.J., Hallmans, G., Hartikainen, A.L., Hayward, C., Hernandez, D., Hicks, A.A., Holm, H., Hung, Y.J., Illig, T., Jones, M.R., Kaleebu, P., Kastelein, J.J.P., Khaw, K.T., Kim, E., Klopp, N., Komulainen, P., Kumari, M., Langenberg, C., Lehtimäki, T., Lin, S.Y., Lindström, J., Loos, R.J.F., Mach, F., McArdle, W.L., Meisinger, C., Mitchell, B.D., Müller, G., Nagaraja, R., Narisu, N., Nieminen, T.V.M., Nsubuga, R.N., Olafsson, I., Ong, K.K., Palotie, A., Papamarkou, T., Pomilla, C., Pouta, A., Rader, D.J., Reilly, M.P., Ridker, P.M., Rivadeneira, F., Rudan, I., Ruokonen, A., Samani, N., Scharnagl, H., Seeley, J., Silander, K., Stancáková, A., Stirrups, K., Swift, A.J., Tiret, L., Uitterlinden, A.G., Van Pelt, L.J., Vedantam, S., Wainwright, N., Wijmenga, C., Wild, S.H., Willemsen, G., Wilsgaard, T., Wilson, J.F., Young, E.H., Zhao, J.H., Adair, L.S., Arveiler, D., Assimes, T.L., Bandinelli, S., Bennett, F., Bochud, M., Boehm, B.O., Boomsma, D.I., Borecki, I.B., Bornstein, S.R., Bovet, P., Burnier, M., Campbell, H., Chakravarti, A., Chambers, J.C., Chen, Y.D.I., Collins, F.S., Cooper, R.S., Danesh, J., Dedoussis, G., De Faire, U., Feranil, A.B., Ferrières, J., Ferrucci, L., Freimer, N.B., Gieger, C., Groop, L.C., Gudnason, V., Gyllensten, U., Hamsten, A., Harris, T.B., Hingorani, A., Hirschhorn, J.N., Hofman, A., Hovingh, G.K., Hsiung, C.A., Humphries, S.E., Hunt, S.C., Hveem, K., Iribarren, C., Järvelin, M.R., Jula, A., Kähönen, M., Kaprio, J., Kesäniemi, A., Kivimaki, M., Kooner, J.S., Koudstaal, P.J., Krauss, R.M., Kuh, D., Kuusisto, J., Kyvik, K.O., Laakso, M., Lakka, T.A., Lind, L., Lindgren, C.M., Martin, N.G., März, W., McCarthy, M.I., McKenzie, C.A., Meneton, P., Metspalu, A., Moilanen, L., Morris, A.D., Munroe, P.B., Njølstad, I., Pedersen, N.L., Power, C., Pramstaller, P.P., Price, J.F., Psaty, B.M., Quertermous, T., Rauramaa, R., Saleheen, D., Salomaa, V., Sanghera, D.K., Saramies, J., Schwarz, P.E.H., Sheu, W.H.H., Shuldiner, A.R., Siegbahn, A., Spector, T.D., Stefansson, K., Strachan, D.P., Tayo, B.O., Tremoli, E., Tuomilehto, J., Uusitupa, M., Van Duijn, C.M., Vollenweider, P., Wallentin, L., Wareham, N.J., Whitfield, J.B., Wolffenbuttel, B.H.R., Ordovas, J.M., Boerwinkle, E., Palmer, C.N.A., Thorsteinsdottir, U., Chasman, D.I., Rotter, J.I., Franks, P.W., Ripatti, S., Cupples, L.A., Sandhu, M.S., Rich, S.S., Boehnke, M., Deloukas, P., Kathiresan, S., Mohlke, K.L., Ingelsson, E., Abecasis, G.R. (2013) Discovery and refinement of loci associated with lipid levels. Nat. Genet. 45, 1274–1285.

45. Wu, Y., Byrne, E.M., Zheng, Z., Kemper, K.E., Yengo, L., Mallett, A.J., Yang, J., Visscher, P.M., Wray, N.R. (2019) Genome-wide association study of medication-use and associated disease in the UK Biobank. Nat. Commun. 10, 1–10.

46. Yamauchi, T., Kamon, J., Waki, H., Terauchi, Y., Kubota, N., Hara, K., Mori, Y., Ide, T., Murakami, K., Tsuboyama-Kasaoka, N., Ezaki, O., Akanuma, Y., Gavrilova, O., Vinson, C., Reitman, M.L., Kagechika, H., Shudo, K., Yoda, M., Nakano, Y., Tobe, K., Nagai, R., Kimura, S., Tomita, M., Froguel, P., Kadowaki, T. (2001) The fat-derived hormone adiponectin reverses insulin resistance associated with both lipoatrophy and obesity. Nat. Med. 7, 941–946.

47. Zhang, X., Cao, Y.J., Zhang, H.Y., Cong, H., Zhang, J. (2019) Associations between ADIPOQ polymorphisms and coronary artery disease: A meta-analysis. BMC Cardiovasc. Disord. 19.

48. Zhao, Z., Xu, D., Wang, Zheng, Wang, L., Han, R., Wang, Zhenzhen, Liao, L., Chen, Y. (2018) Hepatic PPARα function is controlled by polyubiquitination and proteasome-mediated degradation through the coordinated actions of PAQR3 and HUWE1. Hepatology 68, 289–303.

